# Real-World Treatment Patterns of Patients with Basal Cell Carcinoma Using Sonidegib and Vismodegib

**DOI:** 10.1101/2025.09.29.25336664

**Authors:** Mark Lebwohl, Darrell Rigel, Zeynep Eroglu, Victoria Barghout, Deepshekhar Gupta, Enrico Zanardo, Lynn Huynh, Mihran Yenikomshian, Nicholas Squittieri, Thomas Ferro, Kunal Patel

**Author notes:** Affiliation at the time of the study (Thomas Ferro). **Corresponding author:** Mark Lebwohl Icahn School of Medicine at Mount Sinai 1 Gustave L Levy Pl New York, NY 10029 **Email:**. **Prior Presentations:** Posters presented at the Academy of Managed Care Pharmacy Nexus 2024, the 44th Annual Fall Clinical Dermatology Conference 2024 (encore), Winter Clinical Dermatology Conference—Miami 2025 (encore), Maui Derm Hawaii 2025 (encore), Winter Clinical Dermatology Conference—Hawaii 2025 (encore), the 2025 American Academy of Dermatology Annual Meeting, and Academy of Managed Care Pharmacy 2025 (encore).

## Abstract

**Introduction:** Sonidegib and vismodegib are Hedgehog pathway inhibitors (HHIs) approved for the treatment of locally advanced basal cell carcinoma (laBCC), as well as metastatic basal cell carcinoma (mBCC) for vismodegib. Few studies have compared real-world treatment patterns associated with HHI treatment.

**Objective:** To investigate the real-world treatment patterns and conditions of patients receiving HHIs for BCC.

**Methods:** In this longitudinal study, claims from the Komodo Health Claims Database (between 2016 and 2023) were used to identify patients. Baseline characteristics and comorbidities of patients were assessed. Time to treatment discontinuation (TTD), odds of discontinuation, and clinical conditions experienced during treatment were analyzed.

**Results:** Patients who received sonidegib remained on treatment longer than those on vismodegib (log-rank test; *P* = 0.041) and were 23% less likely (*P* = 0.036) and 32% less likely (*P* = 0.013) to discontinue treatment at 6 and 9 months, respectively. Sonidegib-treated patients were less likely to experience gastrointestinal-related conditions (33% less likely; *P* = 0.045), taste and smell-related conditions (71% less likely; *P* = 0.048), and muscle spasms (52% less likely; *P* = 0.009) during treatment compared with patients who received vismodegib.

**Conclusion:** In the real-world setting, sonidegib-treated patients remained on treatment longer than vismodegib-treated patients and were less likely to experience pharmacologically relevant clinical conditions.

**Key Summary Points:**

- Sonidegib and vismodegib are Hedgehog pathway inhibitors approved for the treatment of locally advanced BCC.
- Few studies to date compare the real-world treatment patterns of sonidegib and vismodegib.
- This real-world, retrospective study assessed the treatment patterns and occurrences of pharmacologically relevant clinical conditions of interest in patients with BCC taking sonidegib or vismodegib.
- In this study, sonidegib-treated patients with BCC remained on treatment longer and experienced less frequent pharmacologic-related clinical conditions than patients taking vismodegib, demonstrating the potential therapeutic advantage of sonidegib.
- The results presented in this study may be used to inform clinicians and treatment providers which Hedgehog pathway inhibitor may be best suited for their patients with BCC to optimize treatment outcomes.

## INTRODUCTION

The incidence of basal cell carcinoma (BCC), the most common skin cancer in the world, is increasing both globally and in the US [1, 2]. For most BCCs, surgery (including standard excision, Mohs micrographic surgery, and curettage and electrodessication) is the primary recommendation for the removal of tumors; radiation therapy, topical treatment, cryotherapy, or photodynamic therapy may also be considered in cases where surgery is not feasible [3, 4]. However, systemic therapy may also be recommended, especially in cases of advanced BCC [3, 4]. Systemic therapies may also be used as adjuvant therapies to surgery (i.e., administered after surgery to reduce the risk of recurrence in high-risk or incompletely excised tumors) or neoadjuvant therapies to surgery (i.e., administered before surgery to shrink locally advanced tumors and improve resectability).

BCC oncogenesis is driven by aberrant Hedgehog signaling originating from mutations in the Smoothened (*SMO*) or Patched (*PTCH*) genes, leading to activation of the pathway and overexpression of effectors such as Glioma-associated oncogene homolog 1 (GLI1) [2, 5]. Hedgehog pathway inhibitors (HHIs) are a class of therapeutics developed to treat advanced BCCs via inhibition of Hedgehog pathway signaling receptors such as the protein Smo [5]. Sonidegib and vismodegib are the only 2 HHIs approved in the US, EU, UK, Switzerland, and Australia for the treatment of locally advanced BCC (laBCC) not amenable to curative surgery or radiation therapy; vismodegib is also approved for the treatment of metastatic BCC (mBCC) [6–14].

Both HHIs demonstrated efficacy in patients with laBCC during pivotal clinical trials [15–18]. At the 39-month analysis of the ERIVANCE trial, the objective response rate (ORR) by investigator review was 60.3% among patients with laBCC receiving vismodegib 150 mg daily, which was unchanged from the ORR of observed in the primary analysis and used as the basis for the US Food and Drug Administration (FDA) approval of vismodegib [17, 19]. At the 42-month analysis of the BOLT trial, patients with laBCC receiving sonidegib 200 mg daily had an ORR of 56.1% by central review and 71.2% by investigator review, which was an improvement relative to the response data from the pivotal 6-month analysis that led to the approval of sonidegib [15, 20–22]. A sensitivity analysis in which the BOLT 42-month outcomes were assessed using ERIVANCE-like criteria was also conducted to adjust for the stringent criteria for laBCC tumor evaluation used in the BOLT and ERIVANCE trials (the modified Response Evaluation Criteria in Solid Tumors [mRECIST] and the composite RECIST endpoints, respectively). By ERIVANCE-like criteria, the ORR for sonidegib 200 mg at 42 months was 60.6% and 74.2% by central and investigator review, respectively [20]. By mRECIST, the complete response (CR) rate for sonidegib 200 mg at 42 months was 4.5% and 9.1% by central and investigator review, respectively; by ERIVANCE-like criteria, the corresponding CR for sonidegib 200 mg was 21.2% and 28.8% [20].

Few studies to date compare real-world HHI treatment persistence, rates of discontinuation, and pharmacologically relevant clinical conditions experienced during treatment in patients with BCC. Of the available studies to date, several are focused on treatment patterns of vismodegib only [23–26], assess small cohort sizes [27–30], or analyze treatment patterns of patients taking HHIs as a single cohort without differentiating between sonidegib and vismodegib [31]. Studies of vismodegib usage in real-world patients report ORRs at 28 to 36 months ranging from 73% to 85%, and median time to treatment discontinuation (TTD) of approximately 6 months [23–26]. Due to the earlier approval of vismodegib vs sonidegib (2012 vs 2015), relevant real-world data are more widely available for vismodegib compared with sonidegib [11, 12]. One long-term safety study of sonidegib estimates a median duration of treatment of 8.8 months; nevertheless, no available studies directly compare sonidegib and vismodegib treatment in large cohorts of patients in the real-world setting [32]. This study investigated the real-world treatment patterns of patients with BCC who received sonidegib or vismodegib and assessed the pharmacologically relevant clinical conditions experienced by these patients during treatment.

## METHODS

### Study Design

This real-world, retrospective, longitudinal cohort study utilized claims from the Komodo Health Claims Database, a database including de-identified medical and pharmacy claims for patients enrolled in a US healthcare plan, with census-level representation of all ages, geographies, incomes, races, ethnicities, and risk pools. Claims filled between 1/1/2016 and 3/31/2023 were used in this study (**Figure 1**). The first HHI claim per patient during the study period was defined as their index date. Patients were adults (≥18 years of age as of the index date) with ≥1 diagnosis of BCC prior to index, ≥2 claims for sonidegib or vismodegib, and ≥3 months of clinical activity or continuous enrollment before and after index.

**Fig. 1.**
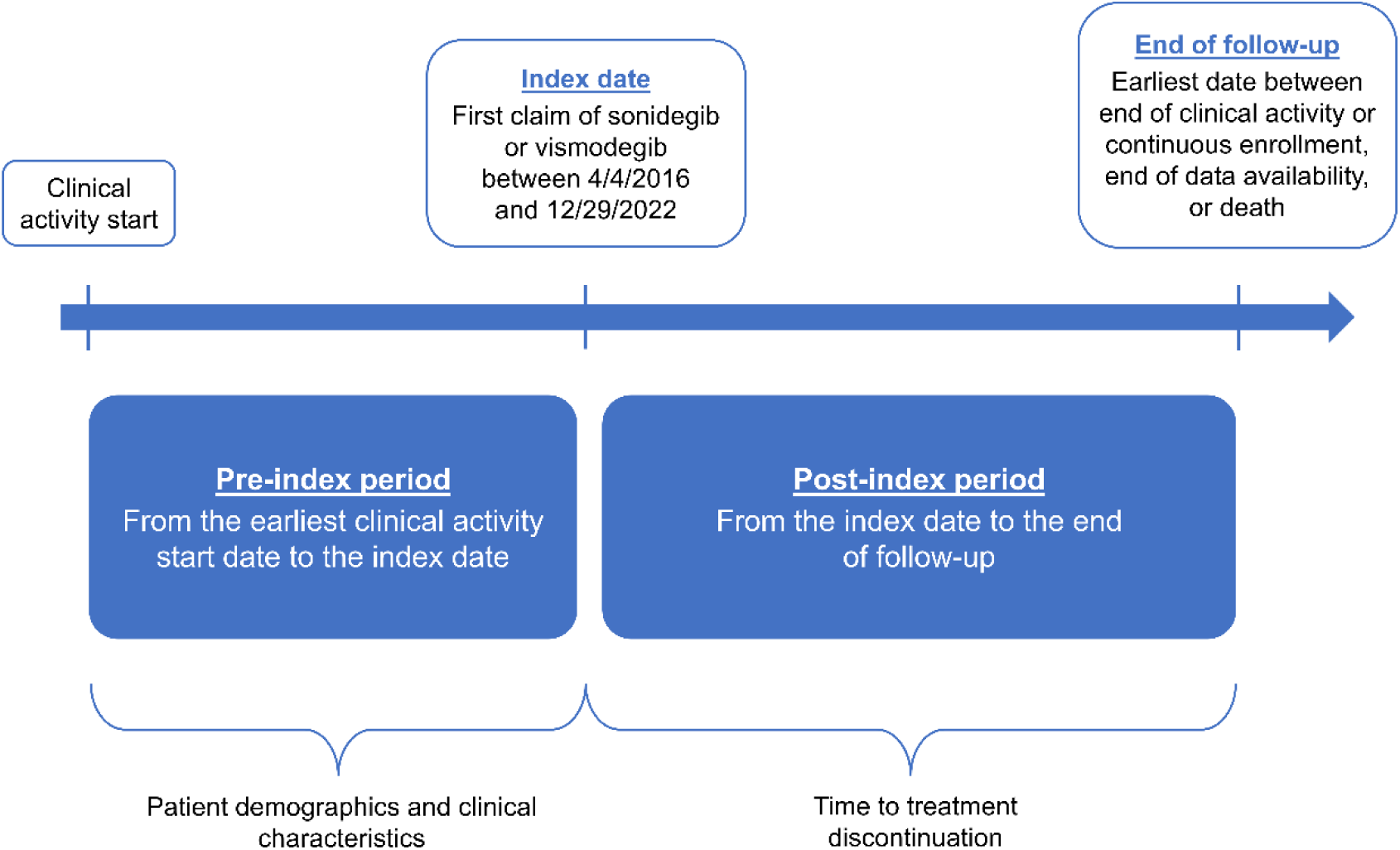
Study design.

The baseline, or pre-index period, was defined as the period from a patient’s beginning of continuous clinical activity or continuous enrollment prior to index until the index date. The follow-up period was defined as the time from index to the earliest of the following: end of clinical activity or continuous enrollment; end of data availability; or death.

### Assessments and Statistical Analyses

Patient demographics were assessed as of the index date, and baseline clinical characteristics (e.g., any comorbidities diagnosed during the pre-index period, location of BCC, and any prior surgery received) were assessed any time prior to the index date. Discontinuation of treatment was defined as a gap with a length twice the FDA-recommended days’ supply of sonidegib (60 days) or vismodegib (56 days) between 2 consecutive claims for index treatment or between the last claim for index treatment and the end of the follow-up period. For patients who discontinued sonidegib or vismodegib, the treatment period was calculated as the time from the index date until the last claim before discontinuation plus 30 or 28 days, respectively. For censored patients, the treatment period was calculated as the time from the index date until end of follow-up. TTD was defined as the time from treatment initiation (or index) to discontinuation or censoring, and was estimated using Kaplan-Meier analyses.

Odds ratios (ORs) of sonidegib vs vismodegib discontinuation at 3, 6, 9, and 12 months were assessed using multivariable logistic regression models. The hazard ratio of discontinuation of sonidegib compared with vismodegib was assessed using multivariable Cox proportional hazards models. All models were adjusted for the following variables: year of index, age at index, specialty of prescribing physician at index, sex, Charlson Comorbidity Index (CCI), geographic region, and surgery 60 days pre- or post-index date (as a proxy for adjuvant or neoadjuvant use of sonidegib and vismodegib). Subgroup analyses were conducted for patients who did not receive surgery 60 days pre- or post-index.

Proportions of patients with a diagnosis (identified using the International Classification of Diseases, Tenth Revision, Clinical Modification [ICD-10-CM] codes) corresponding to the following groups of pharmacologically relevant clinical conditions during treatment were described: gastrointestinal-related conditions, defined as nausea, diarrhea, and weight loss; taste- or smell-related conditions, defined as loss of taste, parageusia, anosmia, and parosmia; muscle-related conditions, defined as malaise and fatigue, weakness and muscle wasting, and muscle spasms; and alopecia. ORs comparing the proportion of patients with a diagnosis corresponding to these clinical conditions of interest during sonidegib vs vismodegib treatment were calculated using multivariable logistic regression models adjusted for the following: year of index, age at index, specialty of prescribing physician at index, sex, CCI, geographic region, surgery 60 days pre- or post-index date, and presence of the clinical condition of interest anytime pre-index. Patients with any diagnosis of alopecia during the pre-index period were excluded from the OR analysis of alopecia during treatment.

## RESULTS

### Patient Demographics and Baseline Clinical Characteristics

Overall, 3766 patients with BCC who received sonidegib (n = 374) or vismodegib (n = 3392) were identified; the median (interquartile range) duration of follow-up was 25.1 (12.3, 42.8) and 29.6 (14.3, 48.6) months for sonidegib- and vismodegib-treated patients, respectively. Most patients were male (sonidegib, 62.8%; vismodegib, 65.0%); the mean patient age was 66.6 years (sonidegib) or 66.1 years (vismodegib; **Table 1**). BCC was located on the face for 40.9% of patients using sonidegib and 47.4% of patients using vismodegib; approximately 20% of sonidegib-treated (19.3%) and vismodegib-treated (18.3%) patients had BCC in multiple locations (**Table 2**). During the pre-index period, patients received destruction surgery (sonidegib, 36.6% and vismodegib, 35.8%), Mohs micrographic surgery (22.7% and 22.7%), malignant surgical excision (22.5% and 21.5%), malignant destruction (19.8% and 18.3%), or reconstructive surgery (0.5% and 0.6%).

**Table 1.**
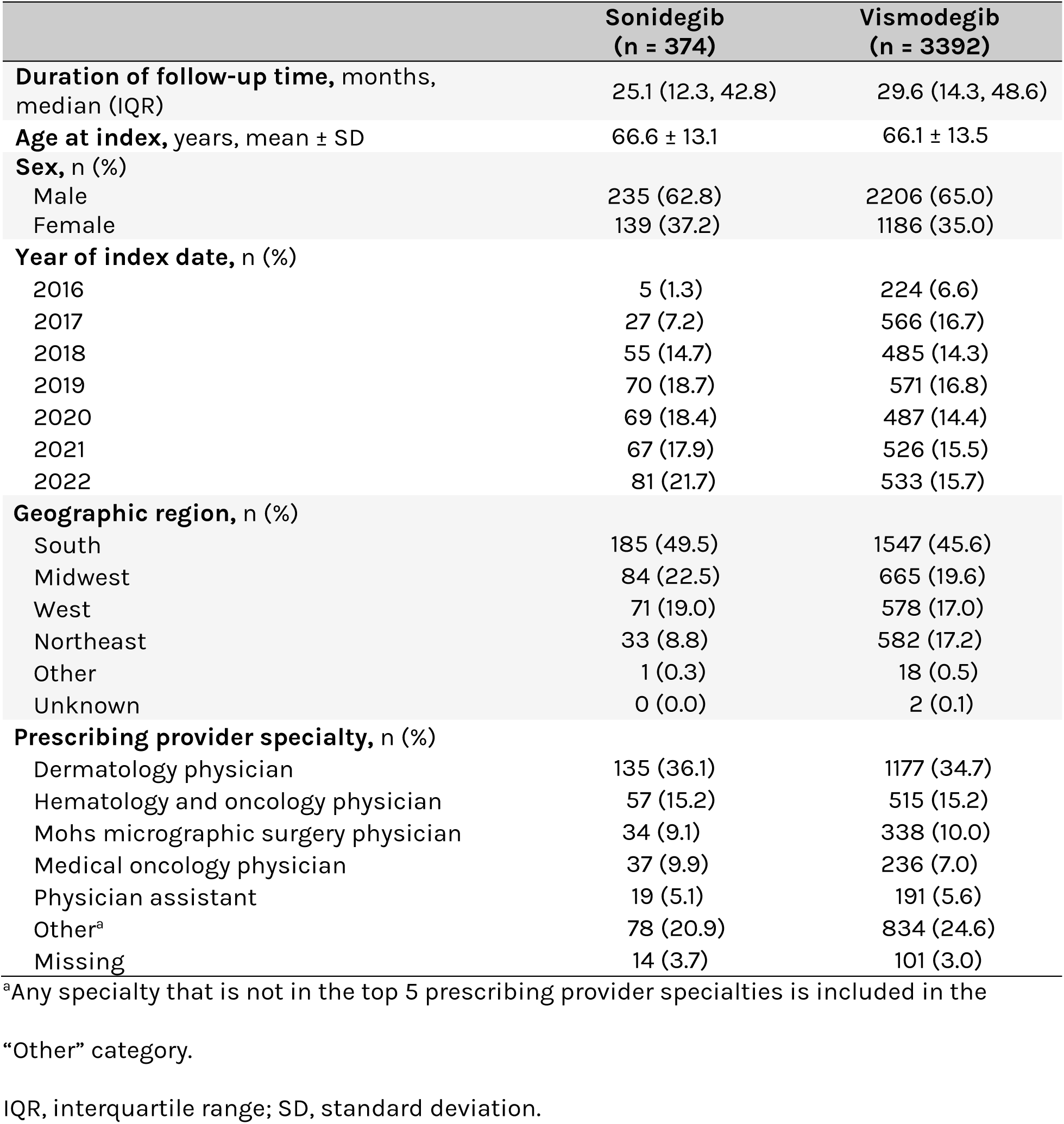
Baseline demographics (on index date) of study population.

|  | Sonidegib<br>(n = 374) | Vismodegib<br>(n = 3392) |
| --- | --- | --- |
| **Duration of follow-up time, months, median (IQR)** | 25.1 (12.3, 42.8) | 29.6 (14.3, 48.6) |
| **Age at index, years, mean ± SD** | 66.6 ± 13.1 | 66.1 ± 13.5 |
| **Sex, n (%)** |  |  |
| Male | 235 (62.8) | 2206 (65.0) |
| Female | 139 (37.2) | 1186 (35.0) |
| **Year of index date, n (%)** |  |  |
| 2016 | 5 (1.3) | 224 (6.6) |
| 2017 | 27 (7.2) | 566 (16.7) |
| 2018 | 55 (14.7) | 485 (14.3) |
| 2019 | 70 (18.7) | 571 (16.8) |
| 2020 | 69 (18.4) | 487 (14.4) |
| 2021 | 67 (17.9) | 526 (15.5) |
| 2022 | 81 (21.7) | 533 (15.7) |
| **Geographic region, n (%)** |  |  |
| South | 185 (49.5) | 1547 (45.6) |
| Midwest | 84 (22.5) | 665 (19.6) |
| West | 71 (19.0) | 578 (17.0) |
| Northeast | 33 (8.8) | 582 (17.2) |
| Other | 1 (0.3) | 18 (0.5) |
| Unknown | 0 (0.0) | 2 (0.1) |
| **Prescribing provider specialty, n (%)** |  |  |
| Dermatology physician | 135 (36.1) | 1177 (34.7) |
| Hematology and oncology physician | 57 (15.2) | 515 (15.2) |
| Mohs micrographic surgery physician | 34 (9.1) | 338 (10.0) |
| Medical oncology physician | 37 (9.9) | 236 (7.0) |
| Physician assistant | 19 (5.1) | 191 (5.6) |
| Other<sup>a</sup> | 78 (20.9) | 834 (24.6) |
| Missing | 14 (3.7) | 101 (3.0) |

<sup>a</sup>Any specialty that is not in the top 5 prescribing provider specialties is included in the
“Other” category.
IQR, interquartile range; SD, standard deviation.

**Table 2.**
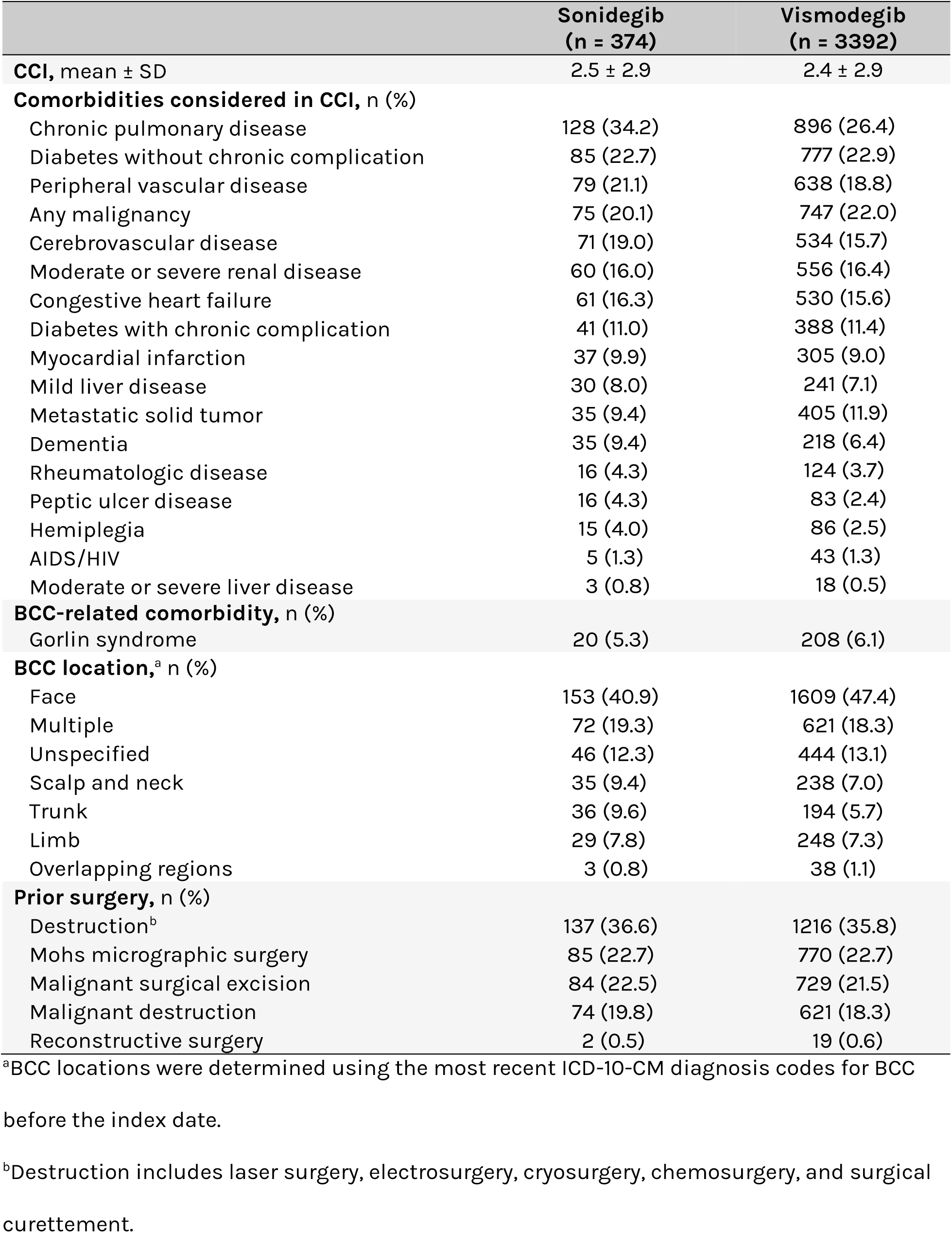
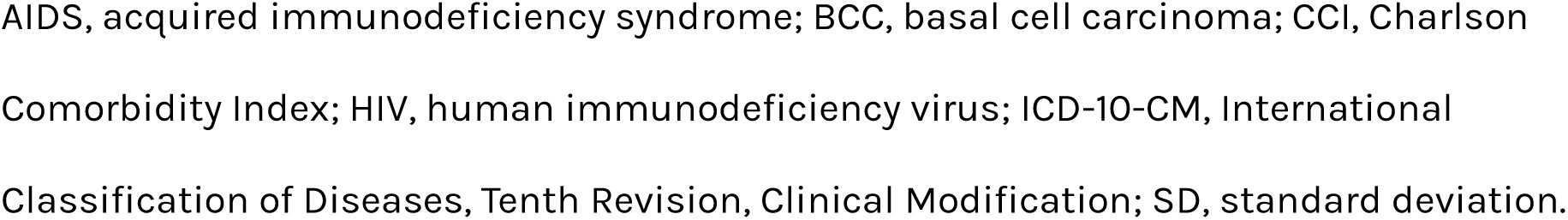
Clinical characteristics and comorbidities (anytime pre-index) of study population.

|  | **Sonidegib**<br>**(n = 374)** | **Vismodegib**<br>**(n = 3392)** |
| --- | --- | --- |
| **CCI, mean ± SD** | 2.5 ± 2.9 | 2.4 ± 2.9 |
| **Comorbidities considered in CCI, n (%)** |  |  |
| Chronic pulmonary disease | 128 (34.2) | 896 (26.4) |
| Diabetes without chronic complication | 85 (22.7) | 777 (22.9) |
| Peripheral vascular disease | 79 (21.1) | 638 (18.8) |
| Any malignancy | 75 (20.1) | 747 (22.0) |
| Cerebrovascular disease | 71 (19.0) | 534 (15.7) |
| Moderate or severe renal disease | 60 (16.0) | 556 (16.4) |
| Congestive heart failure | 61 (16.3) | 530 (15.6) |
| Diabetes with chronic complication | 41 (11.0) | 388 (11.4) |
| Myocardial infarction | 37 (9.9) | 305 (9.0) |
| Mild liver disease | 30 (8.0) | 241 (7.1) |
| Metastatic solid tumor | 35 (9.4) | 405 (11.9) |
| Dementia | 35 (9.4) | 218 (6.4) |
| Rheumatologic disease | 16 (4.3) | 124 (3.7) |
| Peptic ulcer disease | 16 (4.3) | 83 (2.4) |
| Hemiplegia | 15 (4.0) | 86 (2.5) |
| AIDS/HIV | 5 (1.3) | 43 (1.3) |
| Moderate or severe liver disease | 3 (0.8) | 18 (0.5) |
| **BCC-related comorbidity, n (%)** |  |  |
| Gorlin syndrome | 20 (5.3) | 208 (6.1) |
| **BCC location,<sup>a</sup> n (%)** |  |  |
| Face | 153 (40.9) | 1609 (47.4) |
| Multiple | 72 (19.3) | 621 (18.3) |
| Unspecified | 46 (12.3) | 444 (13.1) |
| Scalp and neck | 35 (9.4) | 238 (7.0) |
| Trunk | 36 (9.6) | 194 (5.7) |
| Limb | 29 (7.8) | 248 (7.3) |
| Overlapping regions | 3 (0.8) | 38 (1.1) |
| **Prior surgery, n (%)** |  |  |
| Destruction<sup>b</sup> | 137 (36.6) | 1216 (35.8) |
| Mohs micrographic surgery | 85 (22.7) | 770 (22.7) |
| Malignant surgical excision | 84 (22.5) | 729 (21.5) |
| Malignant destruction | 74 (19.8) | 621 (18.3) |
| Reconstructive surgery | 2 (0.5) | 19 (0.6) |

<sup>a</sup>BCC locations were determined using the most recent ICD-10-CM diagnosis codes for BCC
before the index date.
<sup>b</sup>Destruction includes laser surgery, electrosurgery, cryosurgery, chemosurgery, and surgical curettement.
AIDS, acquired immunodeficiency syndrome; BCC, basal cell carcinoma; CCI, Charlson Comorbidity Index; HIV, human immunodeficiency virus; ICD-10-CM, International Classification of Diseases, Tenth Revision, Clinical Modification; SD, standard deviation.

### Time to Discontinuation

The median TTD was 138 days (4.5 months) for sonidegib and 127 days (4.2 months) for vismodegib. Kaplan-Meier estimates of TTD demonstrate that patients remain on treatment significantly longer (log-rank test, *P* = 0.041) on sonidegib than they do on vismodegib (**Figure 2**). Among patients who did not receive surgery in the 60 days pre- or post-index, the median TTD was 141 days (4.6 months) for sonidegib and 132 days (4.3 months) for vismodegib. TTD in this subgroup was significantly longer (*P* = 0.024) in patients treated with sonidegib compared with vismodegib (**Figure 3**).

**Fig. 2.**
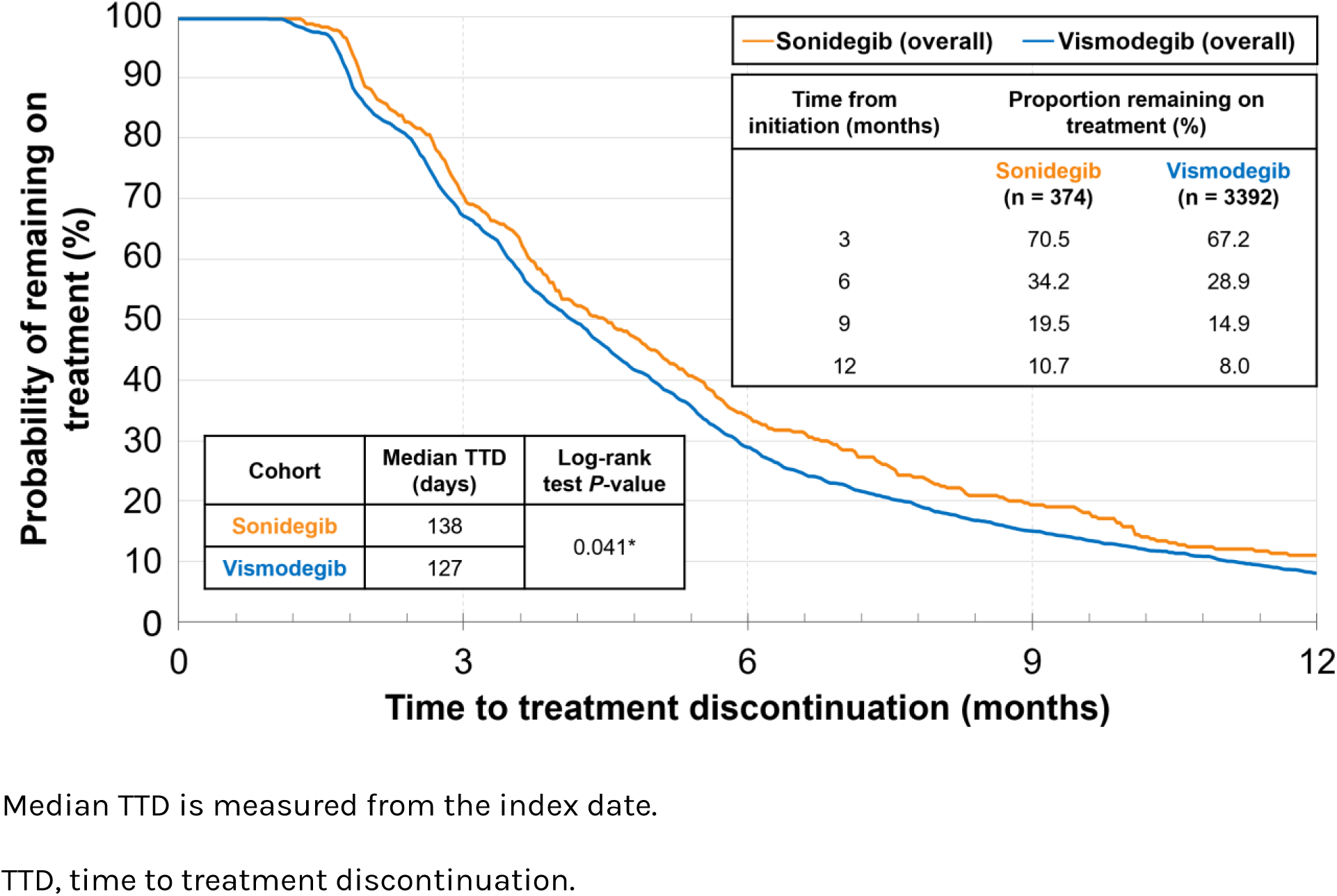
Kaplan-Meier time to treatment discontinuation.

**Fig. 3.**
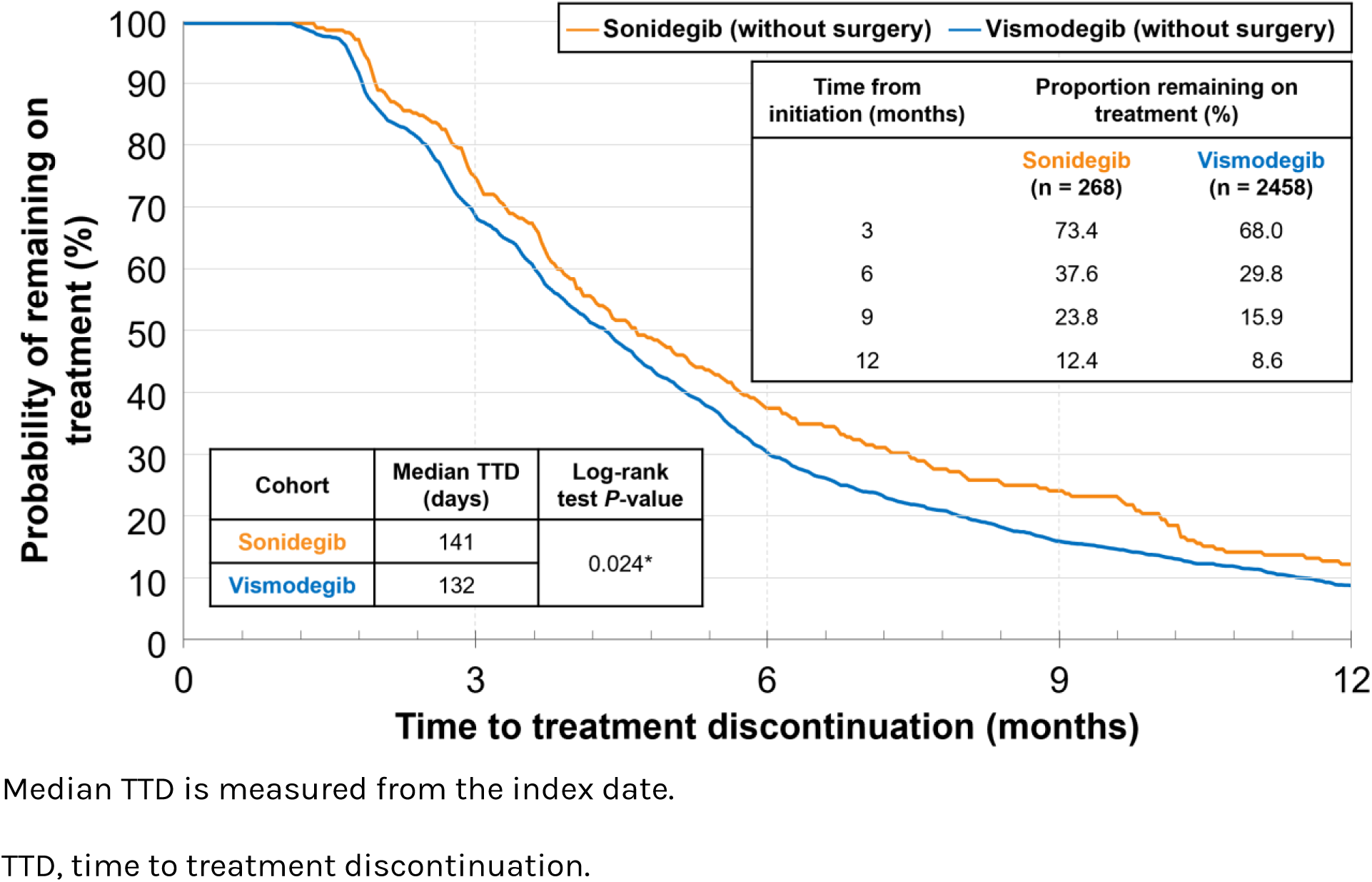
Kaplan-Meier time to treatment discontinuation among patients without surgery.

### Odds of Discontinuation

After adjustment, sonidegib-treated patients were 23% less likely (*P* = 0.036) and 32% less likely (*P* = 0.013) to discontinue treatment at 6 and 9 months, respectively, compared with vismodegib-treated patients (**Table 3**). Among the subgroup of patients without surgery in the 60 days pre- or post-index, sonidegib-treated patients were 29% (*P* = 0.016), 44% (*P* = 0.001), and 39% (*P* = 0.036) less likely to discontinue treatment at 6, 9, and 12 months, respectively, compared with vismodegib-treated patients after adjustment.

**Table 3.** Relative odds of treatment discontinuation of sonidegib vs vismodegib in the overall patient population and in patients without surgery 60 days before or after index.

|  | Total population |  |  | Without surgery |  |  |
| --- | --- | --- | --- | --- | --- | --- |
|  | aOR | (95% CI) | P-value | aOR | (95% CI) | P-value |
| **3 months** | 0.86 | (0.68, 1.10) | 0.266 | 0.76 | (0.57, 1.02) | 0.069 |
| **6 months** | 0.77 | (0.61, 0.98) | 0.036 | 0.71 | (0.53, 0.94) | 0.016 |
| **9 months** | 0.68 | (0.50, 0.92) | 0.013 | 0.56 | (0.40, 0.79) | 0.001 |
| **12 months** | 0.69 | (0.46, 1.05) | 0.086 | 0.61 | (0.38, 0.97) | 0.036 |

Adjusted odds ratios and *P*-values were generated from logistic regression models adjusted for year of index, age at index, physician specialty of prescribing physician at index, sex, CCI anytime pre-index, and geographic region. The overall group was also adjusted for surgery 60 days pre- or post-index date.
aOR, adjusted odds ratio; CCI, Charlson Comorbidity Index; CI, confidence interval.

### Hazard of Discontinuation

The adjusted hazard ratio (aHR) of discontinuation of sonidegib vs vismodegib was 0.89 (*P* = 0.037), indicating that sonidegib-treated patients had an 11% lower rate of discontinuation compared with vismodegib-treated patients. Among the subgroup of patients without surgery in the 60 days pre- or post-index, sonidegib-treated patients had a 14% (aHR = 0.86; *P* = 0.030) lower rate of treatment discontinuation compared with vismodegib-treated patients after adjustment.

### Pharmacologically Relevant Clinical Conditions During Treatment

**Table 4** includes the proportions of patients who experienced specific pharmacologically relevant clinical conditions during treatment. The proportions of patients treated with sonidegib and vismodegib who experienced gastrointestinal-related conditions were 8.3% and 11.9%; taste- or smell-related conditions, 0.8% and 2.1%; muscle-related conditions, 21.1% and 21.2% (muscle spasms, 4.0% and 6.9%); and alopecia, 1.9% and 2.0%, respectively. After adjustment, sonidegib-treated patients were 33% (*P* = 0.045), 71% (*P* = 0.048), and 52% (*P* = 0.009) less likely than vismodegib-treated patients to experience gastrointestinal-related conditions, taste or smell-related conditions, and muscle spasms during treatment, respectively (**Table 5**). In addition, sonidegib-treated patients were 12% (*P* = 0.772) and 8% (*P* = 0.580) numerically less likely to experience alopecia and muscle-related conditions, respectively, during treatment compared with vismodegib-treated patients after adjustment, though this difference was not statistically significant.

**Table 4.** Pharmacologically relevant clinical conditions during treatment.

|  | Sonidegib | Vismodegib |
| --- | --- | --- |
| All patients, n | n = 374 | n = 3392 |
| **Any clinical condition, n (%)** | 102 (27.3) | 977 (28.8) |
| Malaise and fatigue | 42 (11.2) | 321 (9.5) |
| Weakness and muscle wasting | 40 (10.7) | 299 (8.8) |
| Nausea | 21 (5.6) | 232 (6.8) |
| Muscle spasms | 15 (4.0) | 234 (6.9) |
| Weight loss | 10 (2.7) | 144 (4.2) |
| Diarrhea | 7 (1.9) | 120 (3.5) |
| Alopecia | 7 (1.9) | 68 (2.0) |
| Parageusia | 2 (0.5) | 52 (1.5) |
| Loss of taste | 1 (0.3) | 19 (0.6) |
| Anosmia | 0 (0.0) | 5 (0.1) |
| Parosmia | 0 (0.0) | 0 (0.0) |

**Table 5.** Relative odds of clinical conditions among sonidegib-vs vismodegib-treated patients.

|  | aOR (95% CI) | *P*-value |
| --- | --- | --- |
| **Gastrointestinal-related conditions** | 0.67 (0.45, 0.99) | 0.045 |
| **Taste- or smell-related conditions** | 0.29 (0.08, 0.99) | 0.048 |
| **Muscle-related conditions** | 0.92 (0.70, 1.22) | 0.580 |
| Muscle spasms | 0.48 (0.28, 0.83) | 0.009 |
| **Alopecia<sup>a</sup>** | 0.88 (0.37, 2.08) | 0.772 |

<sup>a</sup>Patients with any diagnosis of alopecia during the pre-index period were excluded from the analysis.
Odds ratios, 95% CIs, and *P*-values were generated from logistic regression models adjusted for year of index, age at index, physician specialty of prescribing physician at index, sex, CCI anytime pre-index, surgery 60 days pre- or post-index date, geographic region, and the presence of the clinical condition before index. Since patients with a diagnosis of alopecia in the pre-index period were excluded from the analysis, presence of alopecia before index was not included as a covariate in the comparison of odds of alopecia during treatment.
aOR, adjusted odds ratio; CCI, Charlson Comorbidity Index; CI, confidence interval.

## DISCUSSION

Patients with BCC treated with sonidegib remained on treatment significantly longer and had lower discontinuation rates than patients treated with vismodegib. Sonidegib-treated patients were also significantly less likely to discontinue treatment compared with vismodegib-treated patients at 6 and 9 months after adjusting for key baseline characteristics. These differences were more pronounced in patients who did not use sonidegib/vismodegib adjuvant or neoadjuvant therapy (i.e., patients without surgery in the 60 days pre- or post-HHI initiation). Since efficacy outcomes were not available in claims data, this study could not evaluate how the shorter treatment duration of vismodegib relative to sonidegib relates to the efficacy of the 2 treatments.

Overall, patients taking sonidegib experienced lower rates of pharmacologically relevant clinical conditions during treatment compared with patients taking vismodegib. After adjusting for baseline characteristics, sonidegib-treated patients were less likely to experience gastrointestinal-related conditions, taste- or smell-related conditions, and muscle spasms during treatment compared with patients taking vismodegib.

The findings of greater persistence and a lower rate of pharmacologically relevant clinical conditions with sonidegib compared to vismodegib in this study are consistent with previous real-world studies. In a retrospective, single-center Swiss study, Grossman et al reported that 67% of sonidegib-treated patients and 87% of vismodegib-treated patients discontinued therapy; the mean duration of continuous treatment was 10 months for sonidegib and 17 months for vismodegib among patients surveyed between October 2013 and October 2021 [28]. In the same study, sonidegib-treated patients experienced lower rates of pharmacologically relevant clinical conditions such as dysgeusia, alopecia, muscle spasms, weight loss, and fatigue than vismodegib-treated patients, although they experienced higher rates of malaise, fatigue, and weakness and muscle wasting. Furthermore, patients who switched from vismodegib to sonidegib also experienced lower rates of pharmacologically relevant conditions after switching [28]. In another single-center US-based study, a numerically lower rate of discontinuation of sonidegib (9%) compared to vismodegib (30%) was also observed during a survey period beginning January 2012 and ending June 2021. Furthermore, a numerically lower proportion of sonidegib-treated patients (24%) required dose reductions due to adverse events than vismodegib-treated patients (59%) [33].

Although direct head-to-head comparisons are lacking, the distinct pharmacokinetic profiles of sonidegib and vismodegib may help explain observed differences in treatment duration and discontinuation rates. Sonidegib exhibits both a substantially longer half-life and a markedly greater volume of distribution compared with vismodegib, suggesting broader tissue penetration [11, 34, 35]. Specifically, vismodegib appears to remain largely confined to the plasma compartment, with a volume of distribution between 16 and 27 L, whereas sonidegib, with a volume of distribution exceeding 9000 L, demonstrates extensive tissue distribution [12, 34].

The broader volume of distribution is supported by data showing that sonidegib concentrations in the skin are approximately 6 times higher than in the plasma [34]. Additionally, preclinical studies demonstrate that sonidegib can cross the blood-brain barrier [35]. These characteristics suggest that sonidegib 200 mg daily may provide more sustained Hedgehog pathway inhibition than vismodegib, potentially due to its prolonged systemic presence and enhanced tissue penetration.

Differences in tolerability may also mediate the relationship between pharmacokinetics and treatment persistence. Clinical side effects such as gastrointestinal disturbances, taste and smell abnormalities, and muscle spasms are common reasons for discontinuation of HHIs [15, 18, 19]. It is plausible that the extensive tissue distribution of sonidegib results in lower drug concentrations in HHI-sensitive tissues, thereby reducing adverse effects and improving tolerability. This improved tolerability could, in turn, support longer treatment durations and lower discontinuation rates, potentially leading to better clinical outcomes.

Patients’ clinical conditions and treatment patterns were inferred from codes included in claims data, which do not ensure treatment was taken as prescribed, and are also subject to coding errors. These limitations are expected to affect both cohorts similarly. As this was a retrospective cohort study of claims data, no conclusions about potential efficacy of either treatment can be drawn. Additionally, caution should be exercised when interpreting the results of comparative observational studies. Residual confounding may occur because certain clinical variables that are associated with outcomes of interest may not have been available in the claims data.

## CONCLUSIONS

The results in this study are among the first to present the treatment patterns of patients who received HHI treatment for BCC in a real-world setting. Overall, patients with BCC treated with sonidegib remained on treatment significantly longer and discontinued treatment at lower rates than patients treated with vismodegib. Sonidegib-treated patients also experienced pharmacologically relevant clinical conditions at lower rates than vismodegib-treated patients. The data presented in this manuscript may help inform evidence-based decision-making of clinicians on the best systemic treatment regimens for patients with BCC to improve treatment persistence and clinical outcomes.

## Medical Writing, Editorial, and Other Assistance

Medical writing and editorial assistance in the preparation of this article were provided by June F. Yang, PhD, of Red Nucleus, under the direction of the authors and funded by Sun Pharma.

## Author Contributions

**Mark Lebwohl:** conception, design, writing—review and editing; **Darrell Rigel:** conception, design, writing—review and editing; **Zeynep Eroglu:** conception, design, writing—review and editing; **Victoria Barghout:** conceptualization, methodology, formal analysis, investigation, writing—review and editing; **Deepshekhar Gupta:** methodology, software, validation, formal analysis, investigation, data curation, writing—review and editing; **Enrico Zanardo:** conceptualization, methodology, validation, formal analysis, investigation, writing—review and editing; **Lynn Huynh:** conceptualization, methodology, validation, formal analysis, investigation, supervision, writing—review and editing; **Mihran Yenikomshian:** conceptualization, methodology, supervision, writing—review and editing; **Nicholas Squittieri:** conceptualization, design, formal analysis, supervision, writing—review and editing; **Thomas Ferro:** conceptualization, design, formal analysis, supervision, writing— review and editing; **Kunal Patel:** conceptualization, design, formal analysis, supervision, writing—review and editing.

## Ethics/Ethical Approval

This study did not require ethics committee approval as it is exempt research under 45 CFR § 46.104(d)(4) since it involves only the secondary use of data that were de-identified in compliance with the Health Insurance Portability and Accountability Act (HIPAA), specifically, 45 CFR § 164.514.

## Data Availability

The data that support the findings of this study are available from Komodo Health. Restrictions apply to the availability of these data, which were used under license for this study.

